# Performance of Cardiac MRI for the Diagnosis of Cardiac Amyloidosis in Patients with Advanced Renal Disease

**DOI:** 10.64898/2026.04.06.26350276

**Authors:** Satya Preetham Gunta, Divyanshu Mohananey, Noelle Garster, Christopher Bennett, Sunaina Kalidindi, Jack Geiger, Solomon Ocran, Ravi Narra, Liisa L. Bergmann, David Lewandowski

## Abstract

**Background:** Cardiac MRI (CMR) is often utilized for patients with suspected cardiac amyloidosis (CA). However, data are lacking for use in patients with advanced renal dysfunction (ARD) (GFR<30 mL/min/1.73 m2, dialysis dependent, or renal transplant). This study evaluates the utility of CMR for diagnosis of CA in this population.

**Methods:** Patients with ARD who underwent CMR in a 3T field for suspicion of CA between 2010 and 2024 at our institution were included. A diagnosis of CA was made if any of the following were present a) PYP scintigraphy grade ≥ 2, b) positive endomyocardial biopsy, or c) positive extracardiac biopsy with clinical features of CA. Two CMR-trained physicians independently assessed T1 relaxation time, ECV, Ti scout, LGE, and overall likelihood of CA.

**Results:** Out of the 65 patients included 14 (22%) had a diagnosis of CA. Although T1 time [1352 (1276-1428) ms] and ECV (40.3% +/-9.1%) were elevated across the cohort, they were significantly higher in patients with CA (p<0.001 for both). Both ECV and T1 time reliably predicted CA (AUC of 0.87 and 0.88 respectively). ECV of ≥45% had 75% sensitivity and 80% specificity for CA. A T1 time ≥ 1390 ms had 75% sensitivity and 85% specificity for CA. LGE was prevalent and was seen in 86% and 84% patients with and without CA respectively. Of the 31 patients deemed to be unlikely CA by a CMR reader, 6% had CA. However, of the 34 patients read as possible/likely CA, only 35% had confirmed CA.

**Conclusions:** In this understudied population of ARD, CMR parametric mapping exhibits high negative predictive value (NPV) for CA and improved positive predictive value (PPV) when higher cutoffs are used for T1 time and ECV. CMR reader overall impression exhibits high NPV but low PPV for CA.

## BACKGROUND

Cardiac amyloidosis (CA) is a progressive and potentially fatal condition that is being diagnosed at increasing frequency due to improved diagnostic tools and greater awareness of the condition^1^. While echocardiography remains first line for screening and diagnosis, echocardiographic features such as left ventricular hypertrophy (LVH) lack specificity for the diagnosis of CA, especially in the presence of renal dysfunction. Cardiac magnetic resonance imaging (CMR) with its unique ability for tissue characterization (using a combination of parametric mapping and inversion recovery sequences for late gadolinium imaging) is valuable in patients with suspected CA based on clinical and echocardiographic features^2^. Although, the sensitivity and specificity of CMR for CA in the general population is high^3^, data are lacking for its use in patients with advanced renal dysfunction due to prior concerns with gadolinium contrast in this population. Moreover, there is significant overlap in CMR characteristics of patients with advanced renal dysfunction and those with CA such as increased left ventricular mass, abnormal parametric mapping and high prevalence of late gadolinium enhancement. ^4^ More recently, with the use of Group II gadolinium-based agents (which have been shown to be safe in patients with renal dysfunction), CMR imaging is increasingly being performed in patients with renal dysfunction and suspected CA^5^. Our aim was to study the CMR characteristics of patients with advance renal dysfunction and suspected CA and to evaluate the diagnostic performance of CMR imaging in this population.

## METHODS

### Study Design

In this single-center retrospective cohort study, we included patients with advanced renal dysfunction who underwent CMR for suspicion of CA between 2010 and 2024 at our institution. Advanced renal dysfunction was defined by the presence of at least one of the following criteria: (i) eGFR < 30 mL/min/m2, (ii) receiving renal replacement therapy, or (iii) history of renal transplantation regardless of eGFR. Exclusion criteria were as follows:

i. Patients with history of obstructive coronary artery disease
ii. Moderate or severe left sided valvular heart disease
iii. Hypertrophic cardiomyopathy
iv. Patients with inaccessible/uninterpretable (due to poor quality) CMR images.
v. Patients without alternative confirmatory testing, i.e. nuclear scintigraphy pyrophosphate (PYP) scan and/or tissue biopsy, were also excluded from the study.

A diagnosis of CA was made if any of the following were present: a) PYP scintigraphy with American society of nuclear cardiology grade ≥ 2 uptake, b) positive endomyocardial biopsy or c) extracardiac biopsy positive for amyloid via Congo-red staining *along with* clinical features consistent with cardiac involvement as determined by consensus between a cardiologist specializing in CA (DL) and amyloid hematologist (RN).Demographic data, comorbidities, and lab results were abstracted from the electronic health record.

Baseline characteristics included age, gender, race, subtype of amyloidosis, chronic kidney disease (CKD) stage/presence of renal transplantation, history of coronary artery disease, history of hypertension, history of myocardial infarction, history of diabetes, history of atrial fibrillation, and presence of low QRS voltage on ECG.

This study was approved by the local institutional review board Image acquisition:

i. All CMR exams were performed in a 3 Tesla field (*Siemens Skyra, Erlangen*). Ten minutes after intravenous administration of gadolinium-based contrast agent (GBCA). specifically gadobutrol (Gadavist) dosed at 0.1 mmol/kg), an inversion time (TI) scout sequence was obtained to determine the nulling point (TInull) for normal myocardium.^6^ Having determined TInull, then magnitude inversion recover (IR) gradient echoand 2D breath-hold phase-sensitive inversion recovery (PSIR) sequences were obtained in conventional short- and long-axis planes. TIs varied between approximately 260-320 ms; in order to ensure standardization of our protocol, any discrepancy between magnitude IR and PSIR sequences in regards to late gadolinium enhancement (LGE) was resolved by using PSIR as “source truth” because this sequence cannot “contrast reverse.” Typical sequence parameters included 1.2 mm x 1.2 mm in-plane resolution, 8 mm slice thickness, TE 1.96 ms and ECG-gated TR. ^7,8^
ii. Native and contrast-enhanced T1 mapping was performed using the SSFP based modified look-locker inversion recovery (MOLLI) sequence. Contrast-enhanced T1 mapping was obtained 15 minutes after gadolinium to allow ECV calculation. The upper limit of normal for native T1 and ECV used were 1300 ms and 30% respectively.
iii. Post-processing was performed on *Circle CVI* (*cvi42, Calgary, Canada)*. Late gadolinium enhancement was not quantified. T1 mapping analysis was performed with automated pixel-wise T1 map generation, and ECV was calculated using native and post-contrast T1 values and concomitant hematocrit.

### Image analysis

For this study, each CMR was reviewed independently by 2 level 3 trained CMR readers (DM, NG, CB) who were blinded to the clinical data. Reported values were used for linear and volumetric measurements such as ventricular volumes, ventricular wall thickness, ventricular ejection fraction, myocardial mass, atrial volumes, and left ventricular stroke volume. Each physician independently viewed and post-processed all sequences and provided an overall likelihood of CA (unlikely, possible, or likely).

### Statistical analysis

Data were assessed for normal distribution using the Shapiro Wilk test. Categorical variables are expressed as percentages and continuous variables as mean±SD or median (interquartile range) for skewed variables. We used the Pearson χ2 test for univariate analysis of categorical variables and the independent samples test for univariate analysis of continuous variables. For non-parametric continuous variables, the Man-Whitney U test was used for univariate analysis. Receiver operating curve (ROC) analyses were performed to evaluate test characteristics. A 2-tailed *P*<0.05 was used to denote statistical significance. Statistical analyses were performed using IBM SPSS Statistics (Version 28.0, Armonk, NY).

## RESULTS

### Baseline characteristics and screening results

Our cohort included a total of n= 381 patients with advanced renal disease underwent CMR at our institution. Of those, 75 were performed due to a suspicion for infiltrative disease and had confirmatory testing for CA. The average age of the population was 64, and 50% of the population was African American. 19 (29%) patients were on hemodialysis at the time of the study and 16 (25%) had undergone prior renal transplantation. After excluding patients with indeterminate presence of cardiac amyloid or those performed on 1.5 T machines, a total of 65 patients were included in the study, of which 14 (22%) were deemed to have CA. CA was confirmed in 5 patients via grade ≥2 PYP scan, 4 were diagnosed via positive endomyocardial biopsy, and 5 were diagnosed via positive extra-cardiac biopsy and clinical evidence of cardiac involvement. Of the 14 patients with CA, 5 were diagnosed with ATTR CA, and 9 were diagnosed with AL CA. Troponin and B-natriuretic peptide levels were elevated in all patients where available. ECG analysis demonstrated that most patients in both groups had normal QRS voltage (66%). Patients with CA tended to have low QRS voltage more frequently (21% vs 15%) and patients without CA tended to have high QRS voltage more frequently (20% vs 7%) however these results were not statistically significant.

### CMR Volumetric analysis

Patients who had CA were noted to have significantly higher left ventricular wall thickness compared to those without (Septal wall 17 vs 13 mm p = <0.001, Posterior wall 14 vs 10 mm p=<0.001). Patients with CA also had smaller indexed left ventricular volumes (67 vs 89 mL p=0.002). Notably there was no significant difference in indexed stroke volumes, LV or RV ejection fraction, indexed left ventricular mass, right ventricular volumes, or left atrial volumes between those with or without CA.

### T1/ECV/LGE results

T1 time [1352 (IQR 1276-1428) ms] and ECV (40.3% ± 9.1%) were elevated across the cohort,. T1 relaxation time was higher in patients with CA compared to those without (1423 (IQR 1375-1475) vs 1338 (IQR 1310-1366), p<0.001). Similarly, ECV was higher in CA patients (49.8 ± 7.5 %vs 37.7 ± 7.8 %, p<0.001). Both ECV and T1 relaxation time reliably predicted CA (AUC of 0.87 and 0.88 respectively). An ECV cut-off of ≥45% had 75% sensitivity and 80% specificity for CA. A T1 relaxation time cut-off of ≥ 1390 ms had 75% sensitivity and 85% specificity for CA. Using a combination of these two cut-off values, we found that all patients with CA had either ECV≥45% and/or T1 relaxation time cut-off of ≥ 1390 ms. LGE was highly prevalent in both groups with no significant between group differences in either the presence of pattern of LGE. A mid myocardial pattern was present in 52% of patients and was the most common LGE pattern in our cohort.

### Overall reader impression and TI scout

Expert CMR readers evaluated each study and labelled each with a global impression of likely, possible, or unlikely for CA. For patients without CA, readers graded 29 as unlikely, 12 as possible, and 10 as likely CA. For patients with CA, readers graded 2 as unlikely, 3 as possible, and 9 as likely. The negative predictive value of an unlikely statement was 94%. However, the positive predictive value of a likely or possible statement was only 34%. Assessment of gadolinium kinetics by TI scout demonstrated poor discriminatory ability for CA with 35% of patients without CA having abnormal T1 scout.

## DISCUSSION

In this cohort of patients with advanced renal disease (eGFR <30 mL/min/1.73m^2^) undergoing CMR for suspected infiltrative cardiomyopathy, we retrospectively examined multiple CMR parameters used to identify CA. We found that expert reader impression had strong rule-out value when CA was deemed unlikely, with a negative predictive value of 94%, but limited positive predictive value (34%) when interpreted as possible or likely. Parametric mapping proved to be useful in diagnosis with high AUC values on ROC analysis. A T1 time >= 1390 ms and ECV >= 45% had high specificity. On the contrary, pattern or burden of LGE and gadolinium kinetics via Ti scout were not helpful discriminators.

This is the first study to evaluate the utility of CMR for the diagnosis of CA in the advanced renal disease population. In the general population CMR, particularly when T1 mapping is utilized, is recognized to have high sensitivity and specificity for the detection of CA^3^. Potential mimickers of the LVH/LGE patterns seen on CMR in CA include chronic CAD, Anderson-Fabry’s disease, hypertrophic cardiomyopathy, and hypertensive heart disease. The use of T1 mapping can help rule out some of these mimickers, as few conditions give such large and diffuse elevations of T1 time and ECV than CA. A large meta-analysis determined that an ideal threshold for ruling-out CA on a 3T machine with T1 time was <1161 ms and ruling in CA was > 1337 ms. The same study identified the ideal ECV threshold for ruling out CA was <27.7% and ruling in was >33.4%^9^. In contrast, our study demonstrated much greater elevation of T1 times and ECV across the cohort with particularly elevated values in patients with CA.

In patients with advanced renal disease myocardial structural and tissue characteristics are substantially altered with left ventricular hypertrophy, diffuse elevation of native T1 times, and LGE noted on CMR^10^. This phenomenon is multifactorial including metabolic derangements and activation of the renin–angiotensin–aldosterone system (RAAS) from increased preload and afterload, promoting cellular hypertrophy and driving interstitial fibrosis^11^. The result is a pattern that can mimic infiltrative disease on CMR. A potential confounder could be that at our institution patients with GFR <30 are given single dose gadolinium, which has been suggested to increase calculated ECV by 0.9%^12^ Given that 0.9% is a relatively small increase compared to the differences we noted between study groups, we believe this is a minimal contributor to our findings.

The findings in our study have potentially substantial clinical impact. When a cardiac MRI indicates the possibility of amyloidosis, confirmatory testing is usually performed with nuclear scintigraphy which can rule in or rule out ATTR CA, and serum blood and urine testing is performed to evaluate for AL amyloidosis. However monoclonal gammopathy of undetermined significance is prevalent in 10-12% of patients with chronic kidney disease.^13^ Hence, uncertainty about the presence of CA created by a false positive CMR often leads to additional testing such as endomyocardial biopsy, exposing patients to risks including pericardial tamponade, arrhythmia, or tricuspid valve injury. Despite these implications, our findings do not suggest that the use of CMR for CA is inaccurate, but rather that it is important that interpretation accounts for the effects of renal disease on the myocardium.

There are several important limitations inherent to our study. This is a single center, retrospective analysis involving a very select subgroup of patients which limits broader generalization of our findings. Because of the select population, the number of confirmed amyloid positive patients is relatively low which limits the ability to perform more advanced statistical analysis. Additionally, only a small percentage of the study population underwent endomyocardial biopsy (EMB). When EMB results were not available determination of the presence or absence of CA was based on an amalgamation of extra cardiac biopsy findings, nuclear scintigraphy, and clinical assessment. Clinical assessment was based on agreement between two amyloid specialists from cardiology and hematology, and objective data was utilized whenever possible to limit heterogeneity. Lastly, most patients had CMR performed on a 3T machine, and thus studies performed on 1.5T machines were excluded.

This study highlights a challenge in the diagnostic assessment of this specific patient population, but requires larger scale studies to confirm our findings and to allow for a deeper analysis on what the ideal cutoffs for mapping parameters are for the identification of CA. The development of PET radiotracers for all subtypes of cardiac amyloidosis may alter the diagnostic algorithms for identifying CA^14^ and understanding the inherent strengths and limitations of CMR for patients with advanced renal dysfunction will be critical in test interpretation.

## CONCLUSION

In this previously understudied population of advanced renal dysfunction, CMR parametric mapping exhibits high negative predictive value (NPV) for CA and improved positive predictive value (PPV) when higher cutoffs are used for T1 time and ECV. CMR reader overall impression exhibits high NPV but low PPV for cardiac amyloidosis in patients with advanced renal disease.

## Data Availability

The data that support the findings of this study are available from the corresponding author upon reasonable request

---

**Figure 1.**
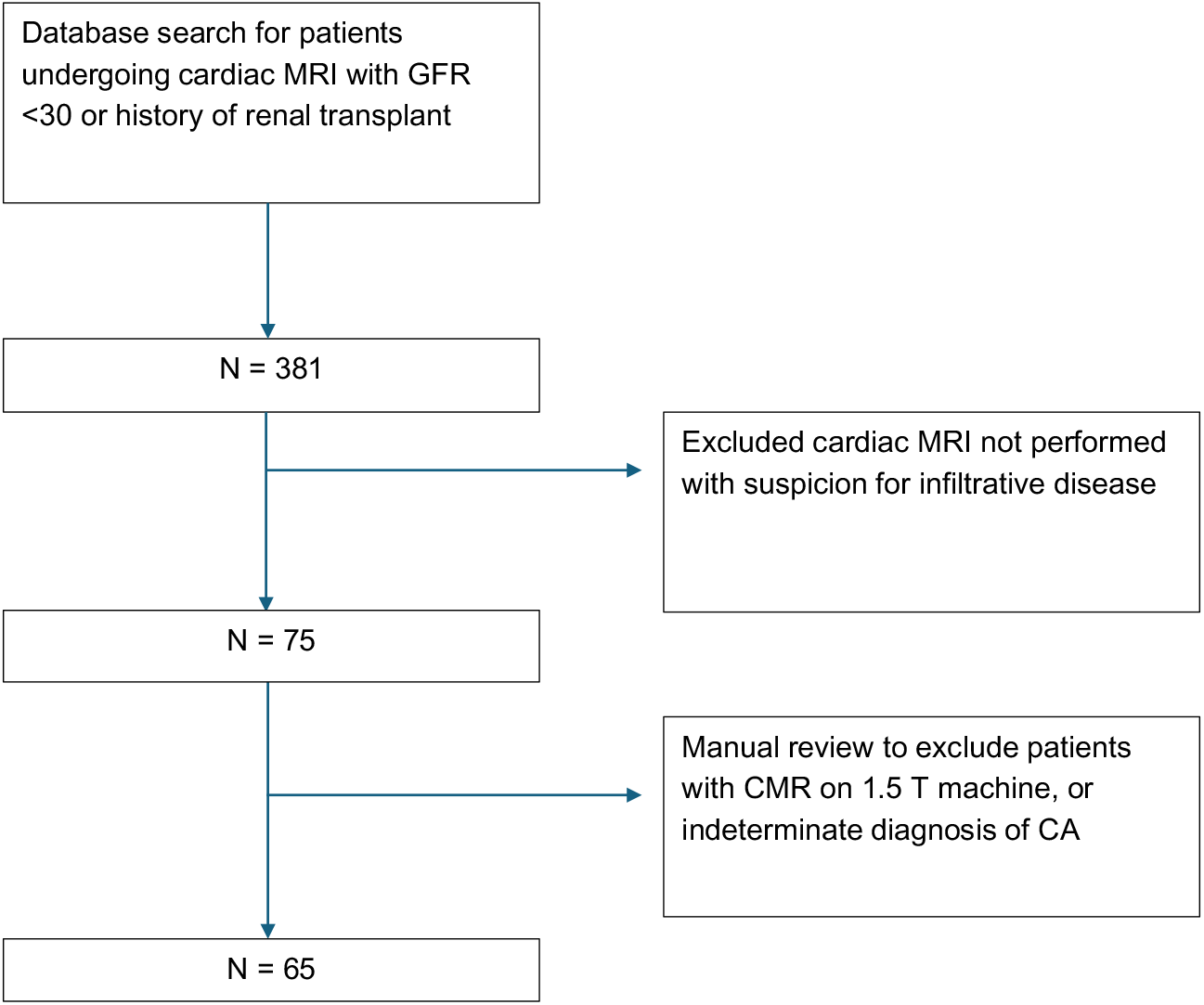
Patient flow chart

**Figure 2.**
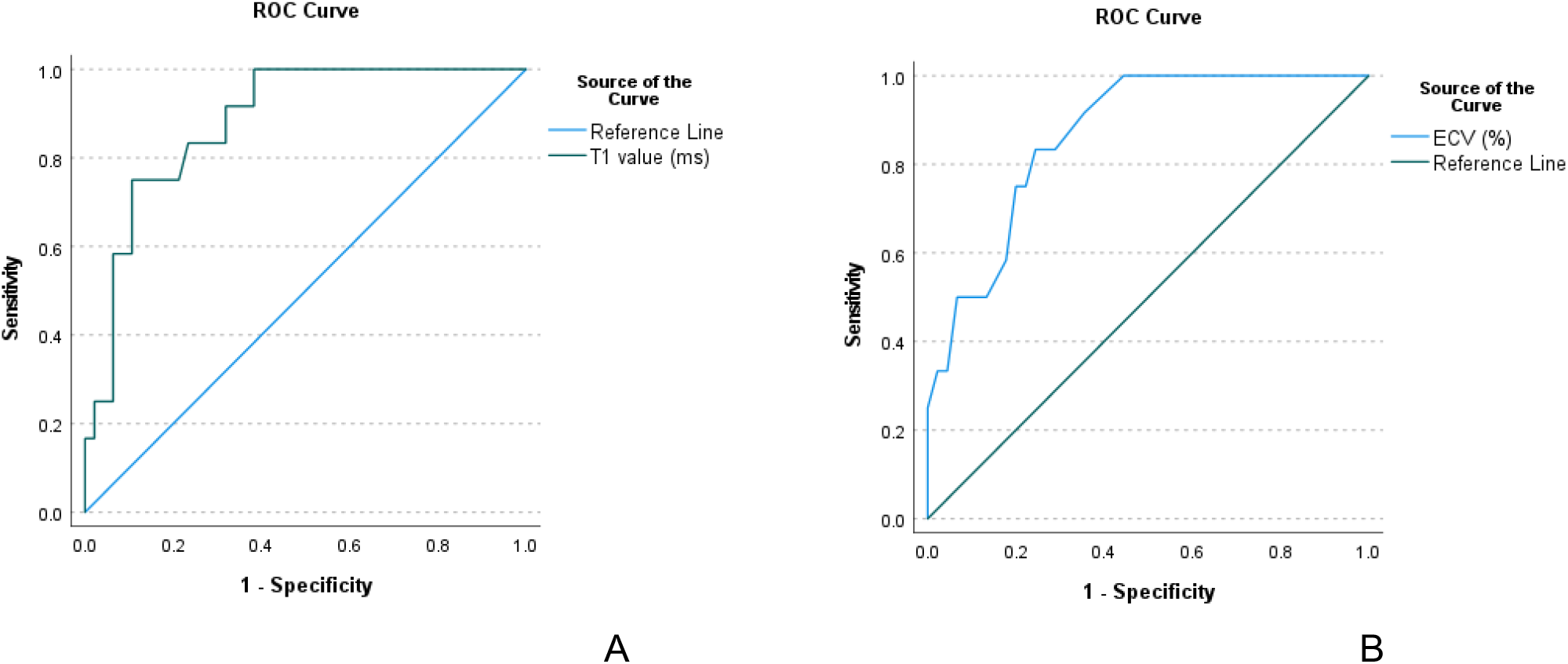
ROC curve analysis for T1 (a) and for ECV (b)

**Figure 3.**
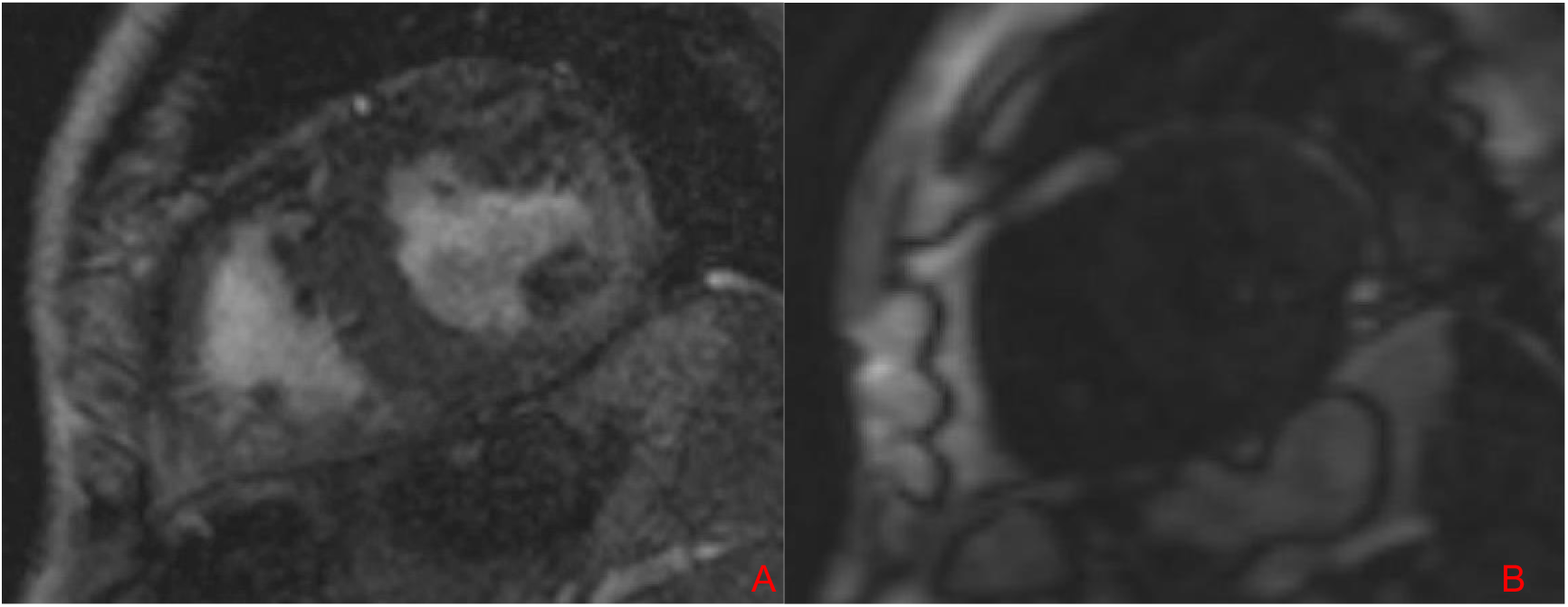
(A) Short axis CMR image demonstrating diffuse late gadolinium enhancement. (B) Ti scout showing simultaneous nulling of myocardium and blood pool. ECG 47% T1 1379 ms. This was ready as likely amyloidosis and ruled out on confirmatory testing.

**Figure 3.**
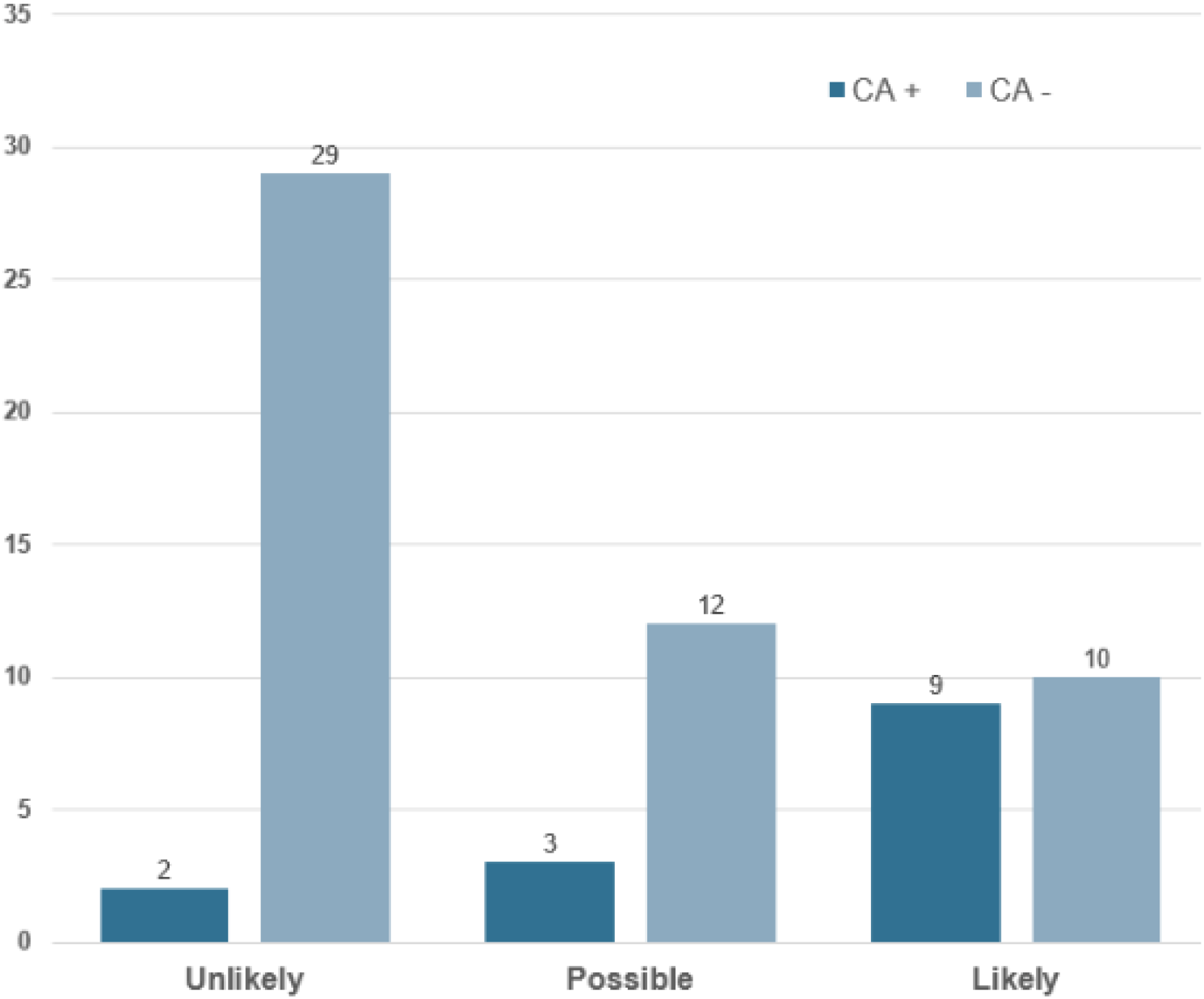
CMR reader interpretation

**Table 1:** Demographic characteristics of included patients.

| Variable | Overall cohort (n=65) | Confirmatory testing positive for cardiac amyloidosis (n=14) | Confirmatory testing negative for cardiac amyloidosis (n=51) | p-value |
| --- | --- | --- | --- | --- |
| Age (years)<sup>a</sup> | 64.1 (13.9) | 70.1 (9.8) | 62.5 (14.4) | 0.067<sup>p</sup> |
| Gender |  |  |  | 0.074<sup>q</sup> |
| Male | 40 (61.5%) | 12 (85.7%) | 28 (54.9%) |  |
| Female | 25 (38.5%) | 2 (14.3%) | 23 (45.1%) |  |
| Race |  |  |  | 0.107<sup>q</sup> |
| African American | 33 (50.8%) | 4 (28.6%) | 29 (56.9%) |  |
| Caucasian | 28 (43.08%) | 8 (57.14%) | 20 (39.22%) |  |
| Other | 4 (6.15%) | 2 (14.29%) | 2 (3.92%) |  |
| CKD |  |  |  | 0.242<sup>q</sup> |
| Stage 4 CKD | 26 (40%) | 8 (57.1%) | 18 (35.3%) |  |
| Stage 5 CKD | 39 (60%) | 6 (42.9%) | 33 (64.7%) |  |
| ESRD on RRT | 19 (29.2%) | 1 (7.1%) | 18 (35.3%) | 0.086<sup>q</sup> |
| h/o Renal transplantation | 16 (24.6%) | 4 (28.6%) | 12 (23.5%) | 0.970<sup>q</sup> |
| h/o Myocardial Infarction | 11 (16.9%) | 2 (14.3%) | 9 (17.6%) | 0.916<sup>q</sup> |
| Coronary artery disease | 35 (53.8%) | 7 (50.0%) | 28 (54.9%) | 0.981<sup>q</sup> |
| h/o CABG | 8 (12.3%) | 1 (7.1%) | 7 (13.7%) | 0.838<sup>q</sup> |
| h/o PCI | 5 (7.7%) | 1 (7.1%) | 4 (7.8%) | 0.632<sup>q</sup> |
| Hypertension | 59 (90.8%) | 12 (85.7%) | 47 (92.2%) | 0.829<sup>q</sup> |
| Diabetes Mellitus (Type I/II) | 40 (61.5%) | 8 (57.1%) | 32 (62.8%) | 0.943<sup>q</sup> |
| Atrial fibrillation | 32 (49.2%) | 7 (50.0%) | 25 (49.0%) | 0.813<sup>q</sup> |
| ECG characteristics |  |  |  | 0.608<sup>q</sup> |
| Normal voltage | 39 (60%) | 10 (71.4%) | 29 (56.9%) |  |
| High voltage | 11 (16.9%) | 1 (7.1%) | 10 (19.6%) |  |
| Low voltage | 11 (16.9%) | 3 (21.4%) | 8 (15.7%) |  |
| Type of Amyloidosis |  |  |  |  |
| AL-Amyloid | 9 (13.8%) | 9 (64.3%) | 0 (0%) |  |
| ATTR-Amyloid | 5 (7.7%) | 5 (35.7%) | 0 (0%) |  |

<sup>a</sup> mean; <sup>p</sup> t-test; <sup>q</sup> Chi square test
CKD: Chronic Kidney Disease; ESRD: End-stage renal disease; RRT: Renal replacement therapy; CABG: Coronary artery bypass grafts; PCI: Percutaneous Coronary Intervention

**Table 2:** MRI characteristics of included patients.

| Variable | Overall cohort (n=65) | Confirmatory testing positive for cardiac amyloidosis (n=14) | Confirmatory testing negative for cardiac amyloidosis (n=51) | p-value |
| --- | --- | --- | --- | --- |
| IVSd (mm) <sup>a</sup> | 13.72 (3.77) | 16.57 (2.766) | 12.92 (3.649) | <0.001 <sup>p</sup> |
| PWd (mm) <sup>a</sup> | 10.89 (3.43) | 13.71 (4.159) | 10.10 (2.762) | <0.001 <sup>p</sup> |
| LV mass indexed to BSA (g/m<sup>2</sup>) <sup>a</sup> | 88.94 (30.62) | 94.93 (24.282) | 87.29 (32.157) | 0.352 <sup>p</sup> |
| LVEDV indexed to BSA (mL/m<sup>2</sup>) <sup>a</sup> | 84.44 (32.189) | 67.36 (16.491) | 89.42 (34.019) | 0.002 <sup>p</sup> |
| LVEF (%) <sup>a</sup> | 54.32(15.777) | 59.86 (16.257) | 52.71 (15.433) | 0.137 <sup>p</sup> |
| Stroke volume indexed to BSA (mL/m<sup>2</sup>) <sup>b</sup> | 39.5 (26) | 34 (15) | 42.5 (29) | 0.212 <sup>r</sup> |
| RVEDV indexed to BSA (mL/m<sup>2</sup>) <sup>b</sup> | 76.5 (44) | 61.5 (47) | 81 (48) | 0.119 <sup>r</sup> |
| RVEF (%) <sup>b</sup> | 53.5 (16) | 54 (36) | 53 (15) | 0.717 <sup>r</sup> |
| LA volume indexed to BSA (mL/m<sup>2</sup>) <sup>a</sup> | 45.56 (17.903) | 43.43 (17.181) | 46.19 (18.237) | 0.590 <sup>p</sup> |
| LGE |  |  |  |  |
| LGE present | 55 (84.62%) | 12 (85.7%) | 43 (84.3%) | 0.898 <sup>q</sup> |
| LGE absent | 10 (15.38%) | 2 (14.29%) | 8 (15.69%) |  |
| subendocardial LGE | 10 (15.38%) | 2 (14.29%) | 8 (15.69%) | 0.258 <sup>q</sup> |
| mid-myocardial LGE | 34 (52.3%) | 5 (35.7%) | 29 (56.9%) |  |
| subepicardial LGE | 4 (6.2%) | 1 (7.9%) | 3 (5.9%) |  |
| mixed pattern LGE | 7 (10.77%) | 4 (28.57%) | 3 (5.88%) |  |
| number of segments <sup>b</sup> | 3 (6) | 8.5 (14.75) | 3 (4.5) | 0.120 <sup>r</sup> |
| Ti scout |  |  |  | 0.210 <sup>q</sup> |
| normal | 38 (58.46%) | 6 (42.86%) | 32 (62.74%) |  |
| abnormal | 25 (38.46%) | 8 (57.14%) | 17 (33.33%) |  |
| T1 value (ms) | 1352 (1323-1396) | 1423 (1375-1475) | 1338 (1310-1366) | <0.001 <sup>p</sup> |
| ECV (%) | 40.3 (9.1) | 49.8 (7.5) | 37.7 (7.8) | <0.001 <sup>p</sup> |
| Degree of suspicion for Amyloidosis based on CMR |  |  |  | 0.003 <sup>q</sup> |
| Unlikely | 31 (47.69%) | 2 (14.29%) | 29 (56.86%) |  |
| Possible | 15 (23.08%) | 3 (21.43%) | 12 (23.53%) |  |
| Likely | 19 (29.23%) | 9 (64.28%) | 10 (19.61%) |  |

<sup>a</sup> mean (Standard Deviation); <sup>b</sup> median (Interquartile range); <sup>p</sup> t-test; <sup>q</sup> Chi square test; <sup>r</sup> Mann-Whitney U test
IVSd: Interventricular septum (diastole); PWd: Posterior Wall (diastole); BSA: Body surface area; LVEDV: Left ventricular end-diastolic volume; LVEF: Left ventricular ejection fraction; RVEDV: Right ventricular end-diastolic volume; RVEF: Right ventricular ejection fraction; LGE: late gadolinium enhancement; ECV: Extracellular volume

